# How healthcare professionals manage uncertainty around hormone replacement therapy (dis-)continuation: the TAPER qualitative Study

**DOI:** 10.64898/2026.10.14.26365532

**Authors:** Sarah Hillman, Laura L. Jones, Nina Kuypers, Eleanor Molloy

**Affiliations:** Department of Applied Health Sciences, School of Health Sciences, University of Birmingham, Edgbaston, Birmingham, B15 2TT; Black Women in Menopause: https://www.blackhealthandbeyond.co.uk/

**Keywords:** Menopause, Hormone Replacement Therapy, Primary Health Care, Health Personnel, Qualitative Research

## Abstract

Many women experience menopausal symptoms after stopping hormone replacement therapy (HRT). An evidence gap remains regarding how women and healthcare professionals (HCPs) make decisions about discontinuing and restarting HRT.

**Aim:** To explore experiences of HCPs that support women when they discontinue and restart HRT.

**Design and setting:** Qualitative study with HCPs working in NHS primary care.

**Method:** ^*^The model describes HCPs’ experiences of supporting women to discontinue HRT as an iterative process characterised by uncertainty. Discussions about discontinuation were commonly prompted by changes in clinical risk or annual HRT review, but decisions about whether and when to stop also depended on women’s symptoms, side effects, life stage and readiness. HCPs described agreeing an individualised discontinuation strategy, providing support during the process, and reviewing women afterwards to consider alternative treatments or, where appropriate, restarting HRT.

Semi-structured interviews with HCPs (*n* = 15) conducted between October 2025 and March 2026. A preliminary coding template was created using the discussion guide as a framework. Each interview was then coded inductively against the template which developed iteratively. Team based discussion helped refine codes into themes and sub-themes, and a subsequent conceptual model was created.

**Results:** The model describes HCPs’ experiences of supporting women to discontinue HRT as an iterative process characterised by uncertainty. Discussions about discontinuation were commonly prompted by changes in clinical risk or annual HRT review, but decisions about whether and when to stop also depended on women’s symptoms, side effects, life stage and readiness. HCPs described agreeing an individualised discontinuation strategy, providing support during the process, and reviewing women after discontinuation to consider alternative treatments or, where appropriate, restarting HRT.

**Conclusion:** HCPs recognise the importance of readiness to discontinue and that decision making should be woman led, with HCP support, clear practice guidance and opportunities to revisit treatment. Further research is needed to support women weigh up their personalised risk and benefit of continuing/discontinuing and identify discontinuation strategies that limit resurgent symptoms.

**How this fits in:**

- Hormone Replacement Therapy (HRT) use has increased substantially, but little is known about how healthcare professionals support women who are considering discontinuation.
- Existing guidance offers limited evidence on when and how HRT should be stopped or how best to manage recurrent symptoms.
- This study shows that discontinuation is an iterative, individualised process shaped by clinical risk, symptoms, life stage and women’s readiness to stop.
- Healthcare professionals would benefit from clearer guidance to support shared decision-making, follow-up and decisions about alternative treatment or restarting HRT.

## Introduction

Hormone replacement therapy (HRT) remains first line treatment for menopausal symptoms(1). Prescribing of HRT has increased substantially in England since 2020 with estimated numbers of patients prescribed HRT increasing by 22%, from 2.3 million in 2022/2023 to 2.8 million in 2024/2025(2). Guidance advises that the indication for HRT is to reduce menopause symptoms and not for reduction in long term health outcomes(3). As such, most women^*^ will at some point stop their HRT. Decisions about when, why and how to stop are multifactorial and decisions are driven by healthcare professional (HCP) recommendations, fear about risks (including cancer), and a preference for a natural approach to menopause(4). Around 40% of women experience resurgent menopausal symptoms upon stop their HRT(5). There is an evidence gap relating to the experiences of women discontinuing HRT and the HCPs advising them, the decisions women and HCPs make when discontinuing and restarting HRT, and the best way to discontinue HRT to minimise resurgent symptoms and the impact on women and health care(6).

The TAPER Study is a mixed methods study comprising of five work packages. The aim of TAPER is to understand the experience of women discontinuing HRT and the HCPs advising them, to establish what women prioritise most when stopping or restarting HRT and conduct a feasibility study. This paper reports the findings and analysis of the interviews undertaken with HCPs working in primary care involved in the care of peri/menopausal women.

## Method

### Study design

TAPER study work package 1 was an exploratory qualitative study, undertaken with HCPs working in primary care within the NHS and providing menopause care (ISRCTN19292146). It is reported in accordance with SRQR(7). In parallel, we interviewed eligible women who had experiences of stopping, attempting to stop, or stopping and restarting HRT, (work under peer review).

Sample size for the interview study was estimated to reach approximately 15-20 HCPs. This remained flexible to provide information power(8)to answer the study question.

### Participant identification and recruitment

HCPs were identified either through the NIHR Research Delivery Network or personal networks. Adverts were shared on social media (LinkedIn). Recruitment for participation was open between October 2025-March 2026. Participants indicated interest in the study by emailing the research team on the project email address. Potential participants were called by one of the study team (SH or EM) on a UK telephone number and sent copies of the consent form and participant information sheet. The telephone call was deemed a necessary step to help eliminate imposter participants(9). Consent forms and participant information sheets were sent as hard copies via post, or online through e-mail, depending on participant preferences.

### Consent procedures

Eligible participants were given time to consider participation and an interview date was scheduled. Consent forms were completed online and returned via e-mail, or as hard copy returned via post, verbal recorded consent was also offered where the consent form statements were read and verbally affirmed by the participant. This recording was held separately to the interview.

### Data collection and analysis

Semi-structured interview guides were developed from the literature including a systematic review published by our group (4) and in collaboration with the TAPER Public and Patient Involvement group (PPI), and the TAPER Project Research Group (PRG). The interview guide can be found (supplement guide 1).

Interviews were held remotely and recorded through the Teams platform, undertaken by SH or EM. At the beginning of each interview HCPs were asked to confirm their job role, how long they have been qualified, their gender and if they considered themselves to be a specialist in women’s health. Audio files were shared securely with a specialist GDPR compliant company for transcription. Transcriptions were cleaned and checked for accuracy and uploaded to NVivo 14 software to support data management and analysis.

A preliminary coding template was created using the discussion guide (supplementary table 1) as a framework to support initial deductive codebook thematic analysis(10). Coding was subsequently undertaken inductively against the framework domains. The framework developed iteratively during the analysis and new codes identified within the data were added. All coding was undertaken by SH and EM. Once all transcripts were coded, SH and EM collectively began to refine codes into themes and sub-themes (supplementary table 2).

Initial themes and sub-themes were discussed with LLJ and NK, and a conceptual model was developed and refined (Figure 1).

**Figure 1.**
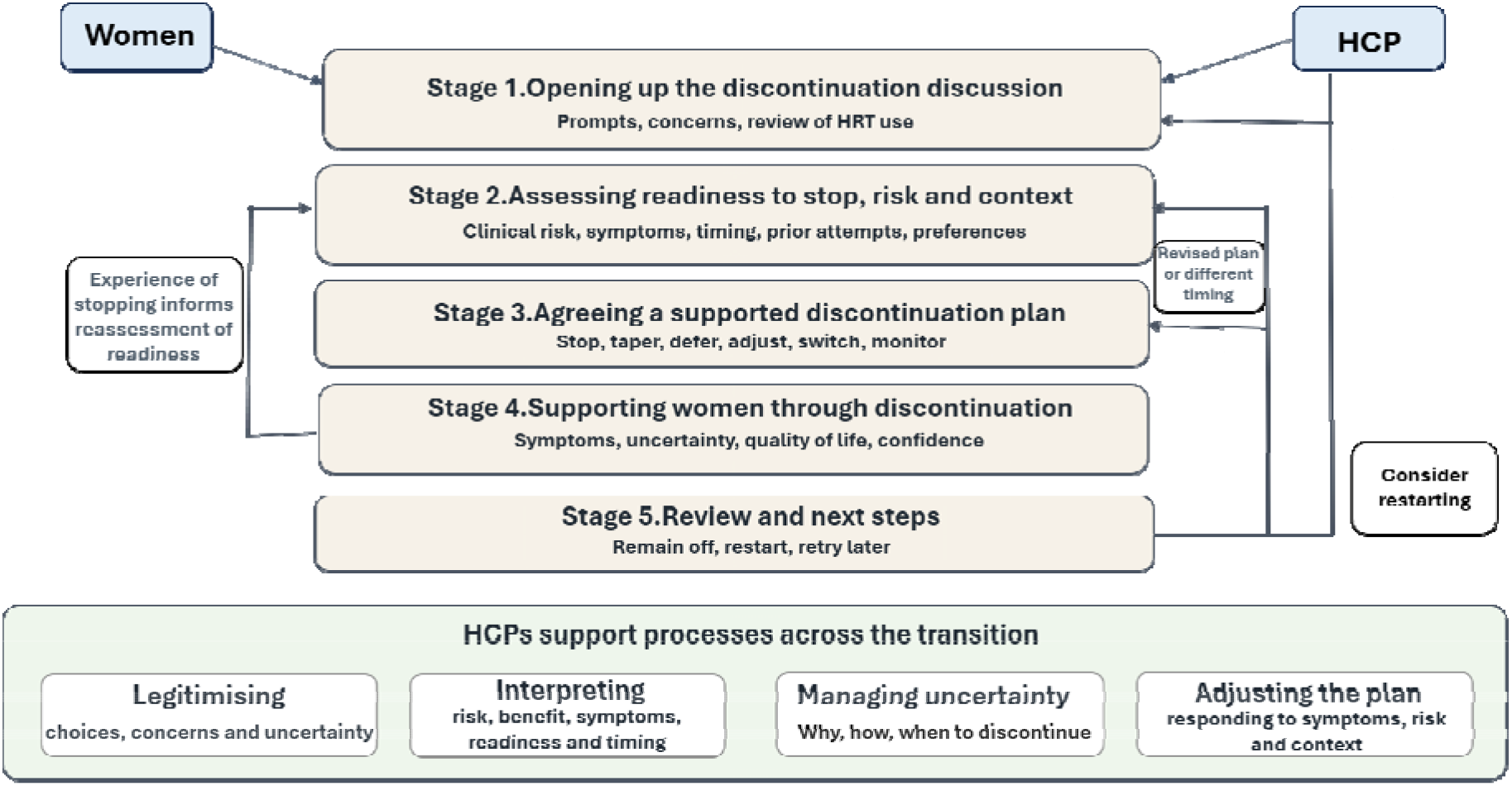

## Ethical approval

Ethical approval was granted by the North-west-Liverpool central Research Ethics Committee 25/NW/0168

## Results

We had initial contact with 20 HCPs. In five cases interviews did not proceed due to the HCP’s capacity, resulting in a total dataset of 15 interviews. All 15 HCPs were female, two participants were South Asian, one was Black and 12 were White. Participants were from varied regions in England with a range of clinical experience (table 1).

**Table 1.** Participant characteristics.

| HCP ID | Job Title | Area | Years since qualified | Special interest in women's health |
| --- | --- | --- | --- | --- |
| 1 | GP | West Midlands | 5 | Yes |
| 2 | GP | North England | 9 | Yes |
| 3 | GP | South East England | 13 | Yes |
| 4 | Advanced nurse practitioner | West Midlands | 27 | Yes |
| 5 | Clinical pharmacist | West Midlands | 15 | Yes |
| 6 | Clinical pharmacist | West Midlands | 3 | No |
| 7 | Advanced nurse practitioner | West Midlands | 12 | Yes |
| 8 | Advanced nurse practitioner | West Midlands | 16 | Yes |
| 9 | GP | West Midlands | 17 | Yes |
| 10 | Clinical Pharmacist | West Midlands | 29 | No |
| 11 | GP | South East England | 24 | No |
| 12 | GP | South West England | 3 | Yes |
| 13 | GP | East Midlands | -Not reported | Yes |
| 14 | GP | North England | 12 | Yes |
| 15 | GP | West Midlands | 16 | No |

The Model is described in detail below and describes how HCPs experience supporting women through the transition of stopping HRT.

**Model for Health Care professionals Figure 1**.

**Supporting HRT discontinuation; an iterative transition with uncertainty**

### Description of the conceptual model

HCPs in primary care described that discussing discontinuation of HRT with women was commonplace either as the focus or as part of a wider consultation about menopause and HRT. They explained that overarching these discussions was;

- the need to **legitimise** women’s choices and concerns around the uncertainty of stopping or continuing HRT
- the need to help women attempt to **interpret** risk and benefit, their current symptom profile (and which symptoms were caused by unmasking menopause symptoms and which were caused by discontinuing the HRT itself) and personal readiness/timing when considering stopping HRT
- that the need to **manage uncertainty** about why, how and when women discontinue HRT was difficult to navigate, and the need to supportively convey this uncertainty
- the need to remain flexible and **adjust plans** to discontinue/continue HRT depending on resurgence or severity of symptoms and when new or cumulative risks present (such as family members being diagnosed with breast cancer, or considering the duration of HRT)

These overarching processes (presented within the green box in the model) cut across the stages of HRT discontinuation presented below.

#### Stage 1. Opening up the conversation

HCPs described conversations about discontinuation being initiated by both women and HCPs. For many it was 50:50 who initiated the conversation, but others preferred to wait for women to start a discontinuation conversation for fear of how it would be received

> *“Only if they ask me. I don’t*… *I think because it often feels quite combative”ID13*

##### 1a Prompt: risk

HCPs found that women brought up the conversation of discontinuing because they felt their risk profile had changed. For some it was a scare of possible breast cancer, for others it was a cardiac event. In some cases, HRT had been discontinued abruptly in hospital and the woman sought advice from the GP on discharge. For other women it was a concern that they should consider stopping at a certain age for fear of cumulative risks.

> *“I think because they’re [women] worried about risks of being on it [HRT] long term, or they’ve heard someone say they shouldn’t be on it past a certain age, so they’ll often come with that agenda”*.*ID15*

##### 1b Prompt: review of HRT use

Many HCPs used a scheduled review of medication to discuss HRT use. For many this was annual. This review may be multidisciplinary, led by clinical pharmacists, physicians associates and doctors. One HCP described proactively sending questionnaires to women which were reviewed by a physician associate and women deemed as higher risk booked for review:

> *“We send out a questionnaire every year to people who are on HRT, and it has*… *because there used to be a lot of people who were not being prescribed appropriately by my colleagues, because they don’t know enough about it really. So, it flags it, and then my*… *we have our physician associates look through them, and then they flag […] then they’ll book that*.*” ID9*

HCPs considered cumulative risks with continued use of HRT and the effect of age. They did this at planned HRT reviews with women which was a prompt to discuss continuation and discontinuation. For HCPs the age at which a woman was considered higher risk for continuation varied from mid to late 50s to 60s.

> *So is there an age, mid to late 50s I would like to think that I would proactively arrange a review, done a review, started talking about it, hearing what the person thinks, and hearing what they’re worried about, hearing where they’re at*.*ID11*

However, there was an overarching acknowledgment that women’s symptoms vary in duration, and HRT review consultations must be individualised. Additionally, women’s assessment of their own personal benefit-risk profile may be at odds with the clinician’s view.

> *“Most women will come off it before the age of 60. But there are a small percentage of women who will continue to have bothersome symptoms and feel that it benefits them in the longer term. So, there’s no set time where I say you have to come off it*.*”ID12*

There was reflection that the thinking about HRT duration has changed and that, with appropriate monitoring, arbitrary age cut-offs for discontinuing are unnecessary

> *“When I first started in 2018, I remember the surgery I worked, it wasn’t here, they used to try and get women off HRT routinely. They used to try and say, “Whoa, she’s 78 why is she still on, we need to try and get her off it*.*” But now I think we don’t tend to have those conversations so much, and we continue as long as women want to take HRT*.*”ID10*

It was at these reviews that HCPs often found that women had stopped their HRT abruptly, without consultation and often in error due to running out of medication. Many of these women did not encounter problems on discontinuation.

> *“But often if I do an opportunistic medication review they might*… *women sometimes say, “Oh I’ve stopped it and I feel okay*.*” Then we just remove it off their prescription” ID3*

#### Stage 2. Assessing readiness to stop

HCPs described needing to assess if a woman felt ready to stop HRT. This depended on; women’s perception of their own risk/benefit from HRT (which may be considered at the time of HRT medication review (1b)), if they were having side effects from HRT, and whether they were at the right time in their menopause transition(11) or personal life to stop HRT. Ultimately conversations and choices were led by the woman.

##### 2a Clinical risk: HCPs

Health care professionals described helping women to weigh up the benefits and risks of HRT in an individualised way. They described how these risks change with age but this should be weighed against symptom control, and that HRT reviews allowed risk factors for disease in later life to be modified (i.e. high blood pressure)

> *“Yeah, I think when we look at a lot of the research over 65, we’re going into the territory of risk might weigh outweigh benefit for patients. But saying that, we look at risk and benefits for that particular patient, so we would modify their risk factors with the patients, we would highlight those, and it’s just the severity of their symptoms as well*.*”ID4*

Some HCPs tried to help women understand risk by comparing the risks in terms of exercise or alcohol consumption

> *“So, whilst numbers are small in terms of increased risk, I often relate it to them what that might be if someone doesn’t exercise, or someone’s drinking a glass of wine a night in terms of risks and averages, so that they have a more informed decision on where they go*.*”ID15*

HCPs described that shared decision making was more challenging when risks were uncertain

> *“But people with a strong family history of breast cancer, so people who are not contraindicated to have, so if there’s not*… *they’ve not got a BRCA gene or something like that, but they’ve got lots of family members with breast cancer, I might suggest to them that perhaps they should look at not being on it for more than five to ten years, depending on the age that they’re on. But that’s a shared decision, it’s not a hard ball answer*.*”ID9*

In some cases, HCPs felt they had reached the limit of their own professional knowledge especially where HRT prescribing could be more complex for some women. Here they sought support at? complex menopause meetings or asking peers online, particularly if they felt women were requesting a riskier option (i.e. oral oestrogen with increased VTE risk)

> *“Somebody who was having an oral and they were having migraines, and I ended up putting the question on the BMS website, just really for my protection more than anything else, because we were trying to persuade her then if she was absolutely adamant, she wanted to continue on it that was fine, but maybe if we could persuade her to have more of a transdermal option that might be better*.*”ID7*

##### 2b Symptoms/side effects

Side effects of HRT were described by HCPS as one of the most common reasons women decided to stop HRT. This might be a problem with the formulation, e.g., patches not sticking, the burden of taking medication or unwanted side effects. One of the most reported side effects encountered by HCPs was unscheduled bleeding. This often led to changes in formulation or further, often repeat, investigations (including scans and hysteroscopies).

> *“But tend to just say to them if we’ve tried multiple formulations, “You’ve got Mirena you’re still bleeding, how*… *do you feel like the HRT has given you enough benefit to put up with this bleeding [loss of sound] or if you want to, and you’ve had a biopsy and that’s been fine, and you feel like your symptoms are really under control and you really want to carry on with HRT then they’re fair enough, you can make that decision*.*”ID2*

##### 2c Timing of discontinuation

HCPs reported that women chose to discontinue when they felt they may no longer need HRT and had become post-menopausal

> *“Like all medication that we prescribe we have to decide do I need this? Is this necessary for me?”ID10*

HCPs discussed with women if they were in the “right place” personally to discontinue (such as recently retired) and described that women often expressed the need to continue while juggling other responsibilities, such as care of elderly parents

> *“I’m looking after an elderly parent, and this just really helps keep my levels at bay*.*” So again it is*… *this is very much improving their quality of life, particularly with their symptoms*.*ID5*

#### Stage 3. Agreeing a supported plan

HCPs felt that it was essential to agree a shared plan about how to discontinue HRT, as without the women’s agreement any plan would be unsuccessful.

> *“I don’t think there are clear guidelines, and I don’t think there’s clear certainty. So I tend to say that I don’t know that we know the best way to do it, and that people are really different in my experience in how they react to that, and that it’s really hard to predict without trying, and that my door is open and they can tell me how things are going along the way, and that the*… *depending on what dose they’re on you could try stepping down a dose, or you can go what people often call ‘cold turkey’*.*”ID11*

HCPs discussed needing to explain to women, that where there were no contraindications restarting was an option. They also discussed swapping formulations to aid the tapering process such as changing oral oestradiol to transdermal.

##### 3a Stop vs Taper

HCPs were often asked by women whether to stop HRT abruptly or taper. They explained that their advice was educated “guess work”. When deciding on a strategy, the initial dose of oestradiol the woman was reducing from (i.e. higher dose maybe harder to abrupt stop) and possibly the age of the woman (i.e. older women may find it easier to abrupt stop being further from their last period) might influence discontinuation approaches. Some HCPs felt that tapering was a gentler approach and more likely to be successful, allowing women control of the process. Other HCPs were unsure and offered both options to women.

HCPs felt both approaches had advantages and disadvantages. Abrupt discontinuation could reduce the burden associated with continued medication use but might result in marked symptom resurgence. They also acknowledged that some women stop HRT without seeking clinical advice, and subsequently inform HCPs at HRT reviews(1b).

> *“There have been women that have gone cold turkey, yeah they have, and some have been fine” ID5*

#### Stage 4. Supporting women during discontinuation

HCPs considered benefits, as described by women, to discontinuation including the financial cost of HRT and that many women want to feel they had “completed” treatment. Resurgent menopausal symptoms during discontinuation were often reported to HCPs. HCPs described finding it hard to determine if menopausal symptoms were due to reducing HRT or unmasking ongoing underlying peri-menopausal symptoms.

> *“If you stop it and you get some usually typical vasomotor symptoms are the most palpable thing, is whether that’s going to be short lived and it’s a physiological response to stopping the treatment, or whether it’s evidence that the underlying menopausal symptoms are unabated and they’re ongoing*.*”ID11*

Vasomotor symptoms were commonly described to HCPs as returning during discontinuation and impacting women. Bleeding could also be a problem.

> *“My advice to her was, “Absolutely fine, you can just stop it, there’s no problems there at all*.*” Thinking in your 70s it would probably be okay, and then she came back to me really, really angry, and it completely destroyed our relationship, because she’d stopped it when she was going on a bus trip to the coast, and she says, ‘I was in my brand new white trousers, and I had a massive bleed and it completely ruined the holiday’*.*”ID14*

#### Stage 5. Review after discontinuation and next steps

HCPs described reviewing women after discontinuing HRT and while some had no symptoms, others commonly described vasomotor and sleep problems. HCPs described helping women consider alternative strategies or restarting HRT.

##### 5a Alternatives and lifestyle

When women had resurgent symptoms, HCPs sometimes considered alternative medications (e.g., clonidine, SSRIs, oxybutynin, Gabapentin), but most HCPs experienced limited success with efficacy

> *“I think I’m always a bit to be honest with SNRIs, oxybutynin and things you’re just like they’re associated with quite a lot of side effects and risks as well. So, I don’t really push them. I definitely offer them, and they’re less effective than HRT” ID2*

Lifestyle changes were often discussed and considered good practice

> *“I think I’d always go through that discussion about healthy diet and exercise, and not drinking, not smoking, sleep, to help improve symptoms generally*.*” ID15*

HCPs described watching and waiting, with the hope that most symptoms should settle.

> *“I warn them that that [resurgent symptoms] might happen, and if they get some mild symptoms that come back maybe just give themselves a little bit of time, like two or three months and see how they feel*.*” ID2*

##### 5b Restarting HRT

HCPs were open to restarting HRT particularly when there were no contraindications to prescribing. They considered there were benefits both in terms of symptoms control and longer-term use such as reducing osteoporosis risk.

Formulation wise HCPs would normally return to what the women was previously prescribed unless discontinuation was because of formulation problems(2b). The exception to this for older women stopping oral oestradiol(2a); here HCPs described recommending transdermal oestrogen as a safer option for venous thromboembolism (VTE) risk.

> *“However, if women are genuinely having sweats, and they stop it and the sweats come flooding back and they want to restart it, and they’re accepting of any of the risks that are within their profile then yeah we’d restart it*.*” ID7*

## Discussion

### Summary

Discussions with women regarding stopping HRT were common in primary care but often shaped by uncertainty about how and when to support stopping. It was acknowledged that these conversations were often led by women and needed to be revisited as risks were cumulative and could change particularly after a diagnosis, or annually.

HCPs acknowledged that readiness to stop was individualised and woman-led. Discontinuation conversations happened more often when women were experiencing side effects or problems on HRT, and at transitional time points in women’s lives such as retirement.

HCPs felt they supported women by helping them plan discontinuation which could be revisited. It was acknowledged that knowing how to stop HRT was uncertain but that many HCPs felt tapering the medication would be “gentler”.

HCPs had experience of women presenting with resurgent symptoms when they discontinued HRT but felt it was difficult to know if these symptoms were temporary, and due to withdrawal of HRT or were unmasking underlying untreated symptoms of menopause. In many cases, and in the absence of contraindications, restarting HRT was common practice. This might be the same formulation that was discontinued or offered an opportunity to discuss a switch to safer preparations. Continued conversations about lifestyle factors were considered good practice alongside alternative pharmacological options.

### Comparison with existing literature

Much existing evidence on HCPs views regarding HRT discontinuation was published in the decade following publication of the Women’s Health Initiative study(12) and our published systematic review showed it was common for HCPs to recommend discontinuation and decrease HRT prescribing in the wake of this study(4). This qualitative data indicates a shift in practice towards a more individualised approach, characterised by shared decision making, and fewer rigid age- or duration-based cut offs. However, many HCPs still appeared to hold broad expectations about the age or circumstances in which, HRT should be discontinued.

Similarly to other published literature HCPs spoke about challenges in approaching discontinuation, particularly if women had not initiated a conversation about stopping HRT(13). Previous literature has also found it is common for women to stop without consultation(14, 15) and due to fear of risk or side effects of HRT(4).

Our 2024 systematic review found that tapering HRT was more commonly recommended than abrupt discontinuation, echoing HCPs’ reflections in the present study that tapering represented a “gentler” approach. The review also found considerable variation in tapering strategies, mirroring the diversity of approaches described by HCPs in our study(4).

### Strengths and limitations

This study provides a contemporary overview of HCPs experiences of supporting women discontinuing HRT in England. Strengths include broad geographical coverage and the inclusion of a range of HCP roles involved in HRT discontinuation within primary care. Patient and public contributors were involved throughout, in study design and refinement of data analysis.

A limitati
on was that only female HCPs were included; which may have restricted the range of perspectives. Although this may reflect the profile of HCPs undertaking much of the women’s health work in primary care. There may also have been an element of selection bias, as those who chose to participate may have a particular interest in, or experience of, HRT discontinuation. The ethnic diversity of participants was limited, the study was not able to explore in depth how cultural and ethnic backgrounds, co-morbidity and disability may shape clinician perspectives on HRT discontinuation and restarting.

The interviews were conducted by SH, a white British female academic GP with a special interest in women’s health, and EM, a white British female senior research fellow. Of the participants, interviewed by SH, two were known to her in a clinical capacity and four were in an academic capacity. One participant interviewed by EM was known to her in an academic capacity.

### Implications for research and practice

With increasing numbers of women initiating HRT, decisions about how and when to discontinue are becoming increasingly important. However, the evidence base for discontinuation of HRT remains limited, creating uncertainty for women and HCPs supporting them. HCPs recognised the importance of women’s readiness to discontinue treatment and the need for decision making to be individualised, woman-led and supported with opportunities to revisit decisions. Further research is needed to support women weighing their personalised benefits and risk of continuing or discontinuing, and to establish effective approaches to discontinuation that minimise the resurgence of symptoms.

#### Additional information

This study/project is funded by an NIHR Advanced fellowship award (NIHR304353).

The views expressed are those of the author(s) and not necessarily those of the NIHR or the Department of Health and Social Care.

## Data Availability

All data produced in the present study are not currently available

## Acknowledgements

The authors would like to acknowledge the time and effort of the participants who contributed to the TAPER Study. They would also like to thank the NIHR RDN for their support with study delivery and the TAPER Steering Group and Project groups for their ongoing support, oversight and insights.

## Notes

### Competing Interest Statement

The authors have declared no competing interest.

### Funding

- NIHR: 304353

