## Supplementary for "How healthcare professionals manage uncertainty around hormone replacement therapy (dis-)continuation: the TAPER qualitative Study"

**Supplementary guide 1 Health care professionals semi structured interviews**

- **Introduce yourself as interviewer – name, role.**
- **If the interview is being held remotely: check technology is working correctly; plans for re-joining if the connection is lost; share a backup phone number; discuss use of video etc.**
- **Thank the participant for agreeing to take part.**
- **Remind participant that the interview can be stopped or paused at any point.**
- **Ensure that the consent form has been completed (if written or electronic) or explain that consent will be taken verbally (and that it will need to be recorded) at the start of the interview.**
- **Ensure completion of demographic questionnaire.**
- **Statement on confidentiality, right to withdraw, what happens to data if they withdraw, recording of the discussion.**
- **Review the purpose of the study in general.**
- **Emphasise the value of their views and opinions – there are no right or wrong answers.**
- **Ask if the participant has any questions before starting the discussion.**
- **Switch on audio recorder/start remote platform recording ask participant to re-confirm consent or take and record formal verbal consent.**
- **Ensure that formal verbal consent, if taken, is in a separate audio file to the main interview recording audio file.**

**Demographic questions:**

**What is your current role?**

**How long since you first qualified?**

**How do you define your gender?**

**Do you consider yourself to have a specialist interest in women's health?**

| Objective | Key discussion points | Example direct questions |
|---|---|---|
| **1. To explore the HCP's experience of prescribing HRT** | Why initiated<br>Length of time using<br>Type of HRT<br>Benefits of HRT<br>Any problems with HRT | Why do you suggest initiating HRT to women?<br>What type of HRT do you prescribe (preparations (i.e. transdermal/oral etc and types of progesterone)?<br>How long do you generally advise women take HRT?<br>Is there any age that you generally suggest women |

| | | |
|---|---|---|
| | | stop? If there is an age, why this age? |
| 2. **Explore the decision to stop HRT** | Concerns about continuation<br>Reasons for stopping<br>Other people involved in decision making | What reason/s may lead you to become concerned about continuing someone's HRT?<br>In what ways are conversations about stopping HRT brought up by women, you or both?<br>In your experience, what are the most common reasons women chose to stop HRT? |
| 3. **Explore how women stop their HRT** | How women stop their HRT<br>If women seek advice about stopping<br>Resurgent symptoms on HRT stopping | Do you generally advise to stop suddenly or taper? Why is this?<br>If you suggest tapering, how do you recommend that this be done?<br>Have you noticed women having symptoms when they stop their HRT?<br>If so what symptoms do women describe, and in your experience how long do these last and how are they managed?<br>Do women describe stopping their HRT or reducing their doses without consultation with a HCPs? |
| 4. **Explore if women Restart HRT** | Experience of Why women restart HRT and if they try stopping again | Do you ever advise restarting HRT? In what circumstances and why?<br>If so, what type of HRT do you restart and is it different to the type they have stopped?<br>When HRT is restarted what do you advise about future discontinuation?<br>Do you ever suggest trying an alternative medication other than HRT? In what circumstances and why? |
| 5. **Explore if women's Quality of life is affected** | The impact of quality of life and responsibilities | In your experience does stopping HRT impact a woman's quality of life? |

| | | |
|---|---|---|
| | | If so in what ways (positively and negatively) and in what ways?<br>Do women suggest it impacts on their responsibilities such as work or caring responsibilities? If so how?<br>In your opinion What are the Pros and cons of stopping HRT? |
| **6. Explore equipoise around a trial of abrupt stop vs tapering** | Views on abrupt stopping vs tapering and thoughts about a future trial | What are your thoughts about whether abruptly stopping or tapering HRT is more likely to be successful in terms of discontinuation?<br>What do you think are the pros and cons of each approach?<br>Do you think abrupt stopping or tapering HRT is more or less likely to affect the resurgence of menopause symptoms? How about specifically vasomotor symptoms?<br>If there was a trial looking at abruptly stopping vs gradually tapering HRT would you be willing to recruit? |

**Closing**

- End of the interview.
- Check/clarify points with the interviewee as necessary – check if the interviewee has any questions.
- Explain where they could find the results of the study (study website).
- Reminder about confidentiality, right to withdraw and snowballing for other participants.
- Explain about provision of voucher as thank you for their time.
- Thank them for their time.

Supplementary table 1 Preliminary coding template

| Name |
|---|
| 1. HCPs views on prescribing HRT |
| perceived benefits of HRT |
| perceived quality of life on initiation of HRT |
| perceived side effects of HRT |
| views about duration including age |
| type or formulation do you prescribe |
| why HCPs suggest initiation of HRT |
| women's understand of HRT |
| 2. HCPs views on stopping HRT |
| HCP concerns around continuation of HRT |
| other people involved in decision making to stop HRT |
| reasons why women stop HRT |
| 3. Explore how women stop HRT |
| seeking advice to stop their HRT |
| advising women to stop their HRT abrupt vs taper |
| effectiveness of abrupt vs taper |
| perceived resurgent symptoms and effect on QOL |
| 4. Explore if women restart HRT |
| advice around discontinuation after restarting |
| alternatives to HRT |

| Name |
| --- |
| advice restarting HRT |
| type or formulation of HRT do you restart |
| 5. Equipoise around an RCT for discontinuation |
| equipoise around abrupt stop vs taper |
| willingness to recruit to a trial |
| 6. Other |
| Going above and beyond |
| expectation of the GP |
| extra qualification |
| job types |
| representation |

**Supplementary Table 2 Themes and sub themes**

- *HCPS views on prescribing HRT*
  - *Perceived benefits of HRT*
  - *Perceived quality of life change on initiation of HRT*
  - *Perceived side effects of HRT*
  - *Views about duration including age*
  - *What type or formulation of HRT HCPs prescribe*
  - *Why HCPs suggest initiation of HRT*
- *HCPs views on stopping HRT*
  - *HCPs concerns around continuation of HRT*
  - *Other people involved in decision making to discontinue HRT*
  - *Reasons why women stop HRT*
- *Exploring how HCPs consider women discontinue HRT*
  - *Advice seeking on HRT discontinuation*
  - *How HCPs advice women to stop HRT*
  - *Perceived resurgent symptoms and effect quality of life*
- *Exploring if women restart their HRT*
  - *Alternatives to HRT*
  - *Advice on restarting HRT*
  - *Type and formulation of HRT restarted*
  - *Advice around discontinuation after restarting*
- *Equipoise around Tapering HRT vs abrupt stopping for successfully discontinuing HRT*
- *Willingness to recruit to a trial*
